# Lesion Symptom Mapping of Domain-Specific Cognitive Impairments using Routine Imaging in Stroke

**DOI:** 10.1101/2021.02.17.21251846

**Authors:** Margaret Jane Moore, Nele Demeyere

## Abstract

**Background and Purpose:** This large-scale lesion-symptom mapping study aimed to investigate the necessary neuro-anatomical substrates of 5 cognitive domains frequently affected post stroke: Language, Attention, Praxis, Number, and Memory. This study aims to demonstrate the validity of using routine clinical brain imaging from a large, real-world patient cohort for lesion-symptom mapping.

**Methods:** Behavioural cognitive screening data from the Oxford Cognitive Screen and routine clinical neuroimaging from 573 acute patients was used in voxel-based lesion-symptom mapping analyses. Patients were classed as impaired or not on each of the subtests within 5 cognitive domains.

**Results:** Distinct patterns of lesion damage were associated with different domains. Language functions were associated with damage to left hemisphere fronto-temporal areas. Visuo-spatial functions were associated with damage to posterior occipital areas (Visual Field) and the right temporo-parietal region (Visual Neglect). Different memory impairments were linked to distinct voxel clusters within the left insular and opercular cortices. Deficits which were not associated with localised voxels (e.g. praxis, executive function) represent distributed functions.

**Conclusion:** The standardised, brief Oxford Cognitive Screen was able to reliably differentiate distinct neural correlates critically involved in supporting domain-specific cognitive abilities. By demonstrating and replicating known brain anatomy correlates within real-life clinical cohorts using routine CT scans, we open up VLSM techniques to a wealth of clinically relevant studies which can capitalise on using existing clinical brain imaging.

## Introduction

Decades of neuropsychological, neurological, and neurophysiological research has indicated that some form of functional specialisation exists within the human brain. From early single case evidence, such as the landmark study dissociating memory components in patient HM^1^, to modern neuroimaging studies^2–6^, previous investigations have reliably identified localised, task-specific neural correlates. Within neurosciences, studies investigating the neural underpinnings of cognition within animal models have provided a basic understanding of the necessary neural substrates of many low-level functions^7,8^. However, the generalisability of these animal models to humans is often limited by small sample sizes, non-naturalistic experimental procedures, and an inherent inability to create adequate proxies of higher-order cognition^9,10^. In contrast, functional imaging techniques struggle to discriminate between activity patterns which are minimally sufficient versus strictly necessary for supporting complex cognitive functions in humans. Similarly, transcranial magnetic simulation studies aiming to identify necessary neural correlates are limited by a comparatively incomplete understanding of the physiological effect of this technique and poor spatial resolution^11^. However, naturally occurring lesions from acquired brain injuries, including stroke, provide an opportunity to reliably identify the human neuroanatomy which is causally related to and strictly necessary for specific higher-order cognitive functions^12,13^.

Though many textbooks refer to likely behavioural consequences of particular stroke types and locations, the evidence base for these relationships often relies on single cases and has not been systematically demonstrated in a single overarching study of acute stroke^14,15^. The majority of previous lesion-mapping studies in stroke have aimed to identify the neural correlates of impairment within a single, specific test such as word reading^16,17^, cancellation tasks^18^, or trail-making tests^19–22^ rather than providing a representative overview of multiple stroke-related cognitive deficits. Conversely, studies which have aimed to identify the correlates of a more diverse range of deficits have been limited by their reliance on language-dependent assessment methods^15,23,24^. For example, Zhao et al.^15^ conducted a large-scale VLSM study (N=410) aiming to identify neural correlates of global cognition using the Montreal Cognitive Assessment (MoCA)^23^. Additionally, the subdomains within MoCA were examined. The MoCA requires verbal responses within the majority of sub-tasks (25 of the 30 points), including those aiming to assess separate domains ^23,25,26^. This language-dependence confound was confirmed in Zhao et al.^15^’s results, as left hemisphere language areas were found to be significantly associated within all investigated cognitive domains including attention, which is well-established to be primarily subserved by right-hemisphere areas^27–29^. Further research is needed to determine the neural correlates of domain-specific post-stroke cognitive impairments using assessment methods unconfounded by language demands.

The purpose of the present investigation was to demonstrate the relationship between stroke locations and specific cognitive domain impairments as measured through a brief domain-specific cognitive screen^24^ designed for stroke and inclusive for patients with aphasia and neglect^26^, in the largest lesion-symptom mapping study of acute post-stroke cognitive impairment (n = 573 vs. n = 410^15^). Voxel-Based Lesion Symptom Mapping (VLSM) provides an effective methodology for identifying the necessary neural correlates supporting human cognitive functions^30^. However, the inferential power of VLSM is often greatly limited by the quality of data considered. Ideally, VLSM analyses should aim to include high-quality neuroimaging and behavioural data from large, representative samples of patients with acute brain injury^31^. Unfortunately, it is often impossible to obtain high-quality MR imaging for acute patients as many have contra-indications for MR or are simply not able to lie still for MRI scanning^32,33^. In addition, the substantially higher costs of MRI scanning mean that CT imaging is the standard pathway in most clinical settings^34^. With respect to timing, neuropsychological studies often require lengthy testing and stroke survivors are often not recruited to partake in research until later in recovery. During this recruitment delay, cortical re-organisation and recovery may occur, weakening the relationship between lesion location and behaviour^31,35,36^. Previous research has strongly suggested that VLSM analyses that do not include both acute neuroimaging and acute behavioural data may not be able to precisely identify neural correlates^31,37^. Additionally, many VLSM studies include only small samples, restricted to patients exhibiting only specific stroke types or locations, or very specific, often milder impairments linked to smaller lesions. This frequent reliance on non-representative samples greatly limits the generalisability of many VLSM studies’ findings. Given these common limitations, new avenues for improving VLSM studies are needed.

UK Clinical Stroke Guidelines^38^ require each suspected stroke patient to complete diagnostic CTs and bedside cognitive assessment within the acute stage. These data offer the potential to investigate the validity of using routine clinical CT scans for VLSM in the context of domain-specific cognitive impairments. Acute clinical CT scans are low-resolution and often have incomplete visualisation of the full extent of stroke damage due to the presence of tissue that has been irreversibly damaged but has not yet been replaced by cerebrospinal fluid^34,39^. This temporal development of lesions may lead to underestimations of the extent of damage, which is generally cited as grounds for avoiding CT data in VLSM analyses. However, it also follows that analyses involving acute CTs are extremely unlikely to yield false positives. Therefore, if the feasibility of using these scans for research can be established, any positive VLSM results yielded by employing clinical CT data would provide strong source evidence in identifying necessary neural correlates.

VLSM has the potential to enhance conclusions on the necessity of underlying functional neuroanatomy from neuroimaging, behavioural, and animal-lesion hypotheses via this human lesion approach. The purpose of the present study is to investigate the validity of using routinely collected clinical neuroimaging data to identify the necessary neural correlates supporting well-known domain-specific cognitive impairments. This question is particularly important to explore as large scale lesion mapping approaches capitalising on routine clinical imaging could significantly expand both the power of future lesion mapping, as well as the clinical representative nature of the samples. This expansion would facilitate more ecologically valid conclusions in a patient group that is characterised by often co-occurring changes in structural integrity as well as grey matter associated with age and vascular disease^40,41^.

### Participants

This investigation presents a retrospective analysis of data collected within the OCS-Tablet, OCS-Recovery, and OCS Care studies (NHS REC references 14/LO/0648, 18/SC/0550, and 12/WM/00335). These studies recruited consecutive stroke survivors during acute hospitalisation (<3 weeks post-stroke), regardless of lesion location or behavioural pathology with minimal exclusion criteria, on their ability to remain alert for 15 minutes. The studies intentionally included participants with communication difficulties and severe cognitive impairments, augmenting the sample’s representativeness. All participants with available OCS data and routine clinical neuroimaging were considered. Patients with no visible lesions or clear evidence of multiple, temporally distinct lesions were excluded.

A total of 573 patients (age= 72(sd=13.2), 43.8% female, 9.7% left-handed) met all inclusion criteria. This sample consisted of a mixed aetiology of ischemic (n=455) and haemorrhagic (n=118) stroke (282 R, 233 L, 58 Bilateral) varying in severity from lacunar strokes to large lesions (average volume = 39.2 (SD=56.2) cm^3^, range= 0.015-444.3 cm^3^). The sample size of each conducted VLSM comparison ranged between 499 and 568, due to varying numbers completing each OCS subtest.

### Neuroimaging Data

Routine post-stroke imaging (497 CT, 67 T2, 2 T1, 7 FLAIR) was employed to quantify lesion anatomy. Lesions were manually delineated in native space using MRIcron (McCausland Center for Brain Imaging, Columbia, SC, USA) by experienced investigators blind to behavioural results. Lesion masks were smoothed at 5mm full width at half maximum in the z-direction^31^, binarized (0.5 threshold), reoriented, and warped into 1×1×1mm stereotaxic space using Statistical Parametric Mapping 12 and Clinical Toolbox^42,43^. All normalised scans and lesions were visually inspected for quality.

### Behavioural Data

Routine post-stroke cognitive assessment data from the Oxford Cognitive Screen^24^ was employed to identify behavioural impairments in 5 core cognitive domains. The OCS is a standardised, stroke-specific clinical cognitive screening tool assessing cognitive functions across: language, attention, memory, praxis, and number processing. Each OCS subtest was designed to assess cognitive functions independently, explicitly minimising language-related and visuo-spatial confounds present in many other screens^26^. OCS is currently used as the standard cognitive screen in >1100 settings, recommended in clinical guidelines^38^, and has 7 published language translations and cultural adaptations.

Patient behavioural impairment was binarily classified based on standardised impairment thresholds for each subtest. The language domain includes subtests on expressive language (Picture Naming), receptive language (Semantics) and Reading (Sentence Reading). The Number domain is composed of a Number Writing Task and a Calculation Task. The memory domain contains sub-tests assessing long-term memory (Orientation), prompted verbal recall (Recall), and unprompted episodic recognition (Recognition). Visuo-spatial abilities are measured both in terms of visual perception (Visual Field Task), as well as in terms of visuo-spatial attention (Cancellation Task). These tests lateralised impairment and are therefore split into two impairment groups, each denoting the presence/absence of right or left-lateralised deficits. Finally, Praxis is measured in a meaningless gestures imitation task and Executive Attention is measured through a non-verbal trail making test.

### VLSM Statistics

Separate VLSM analyses were conducted to determine the neural correlates of impairment within each behavioural comparison groups. These VLSM analyses were conducted on a theory-blind voxel-wise basis using NiiStat (https://github.com/neurolabusc/NiiStat). Only voxels with an overlap of ≥5 patients were considered. Given that binarized behavioural data were used, the non-parametric Leibermeister measure was used to evaluate significance. All comparisons were adjusted for multiple comparisons using a highly conservative Bonferroni correction (alpha=0.05). Bonferroni corrections are substantially more conservative than more traditional, alternative VLSM corrections and this was employed to reduce the risk of false positives. Relevant z-statistic significance cut-offs are reported for each test, with negative z-scores representing some association with behavioural impairment. Lesion anatomy was evaluated versus the Harvard-Oxford Cortical^44^ and John’s Hopkins University White Matter atlases^45,46^. All z-statistic maps are available on the Open Science Framework^47^ (https://osf.io/dc7ay/).

### Results

Figure 1 illustrates the lesion overlays for all 573 participants included in this investigation. The highest lesion overlap (n=88) was present within the MCA territory in line with a clinical picture of majority MCA^48^. However, given our large sample size, territories which were affected to a lesser extent still contained sufficient power to detect results (overlap=5-31 patients). Figure 2 provides the visualisation of all lesion overlays for patients exhibiting impairment within each behavioural comparison group.

**Figure 1:**
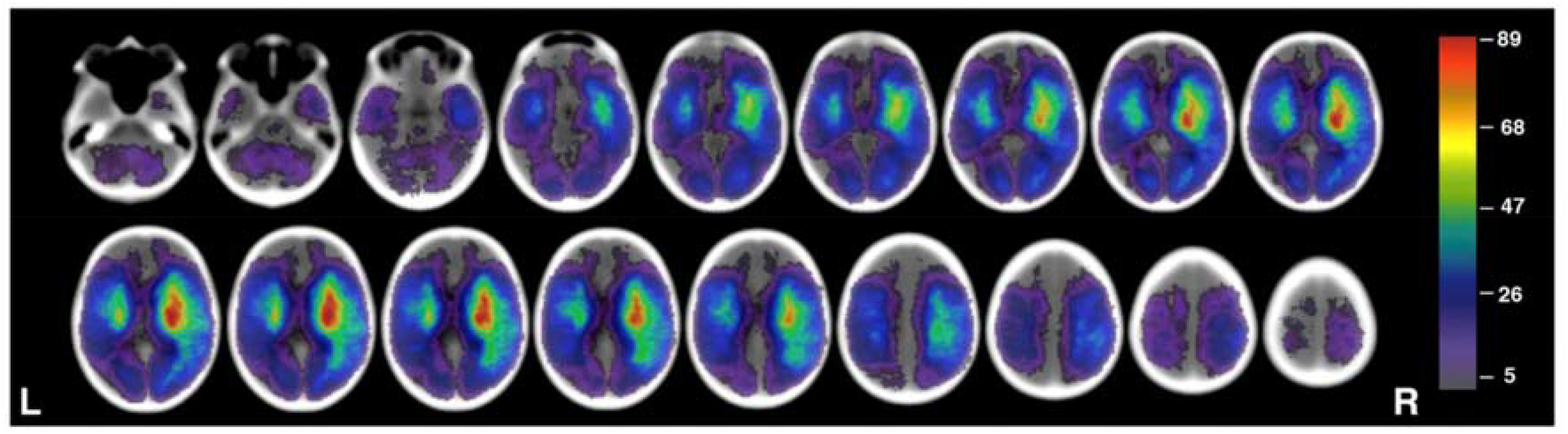
Lesion overlay for all 573 patients in this investigation. Colour represents number of lesions overlapping. Only regions with ≥5 lesions are visualised. Slices within the MNI z-coordinates −44 to +66 are visualised.

**Figure 2:**
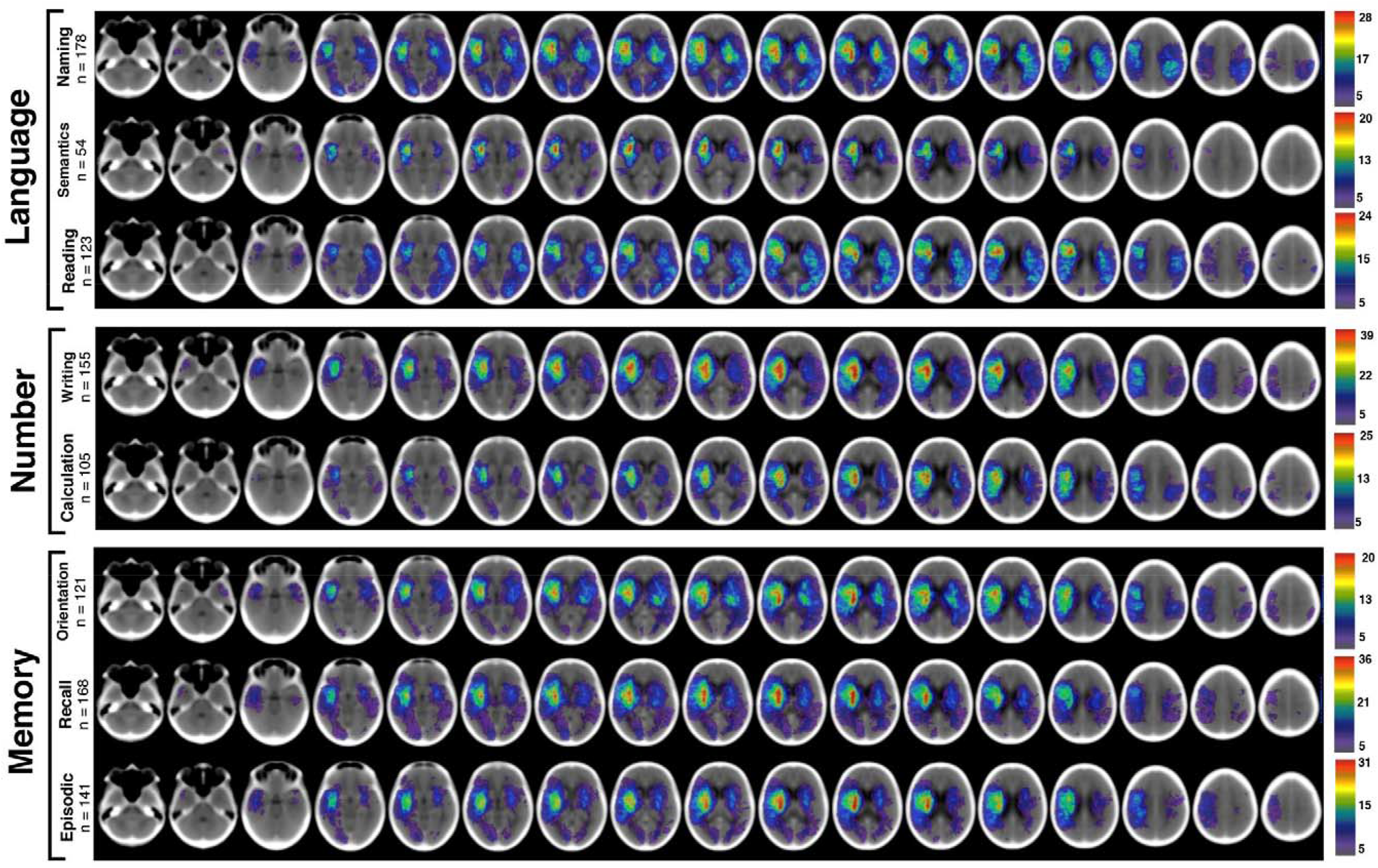
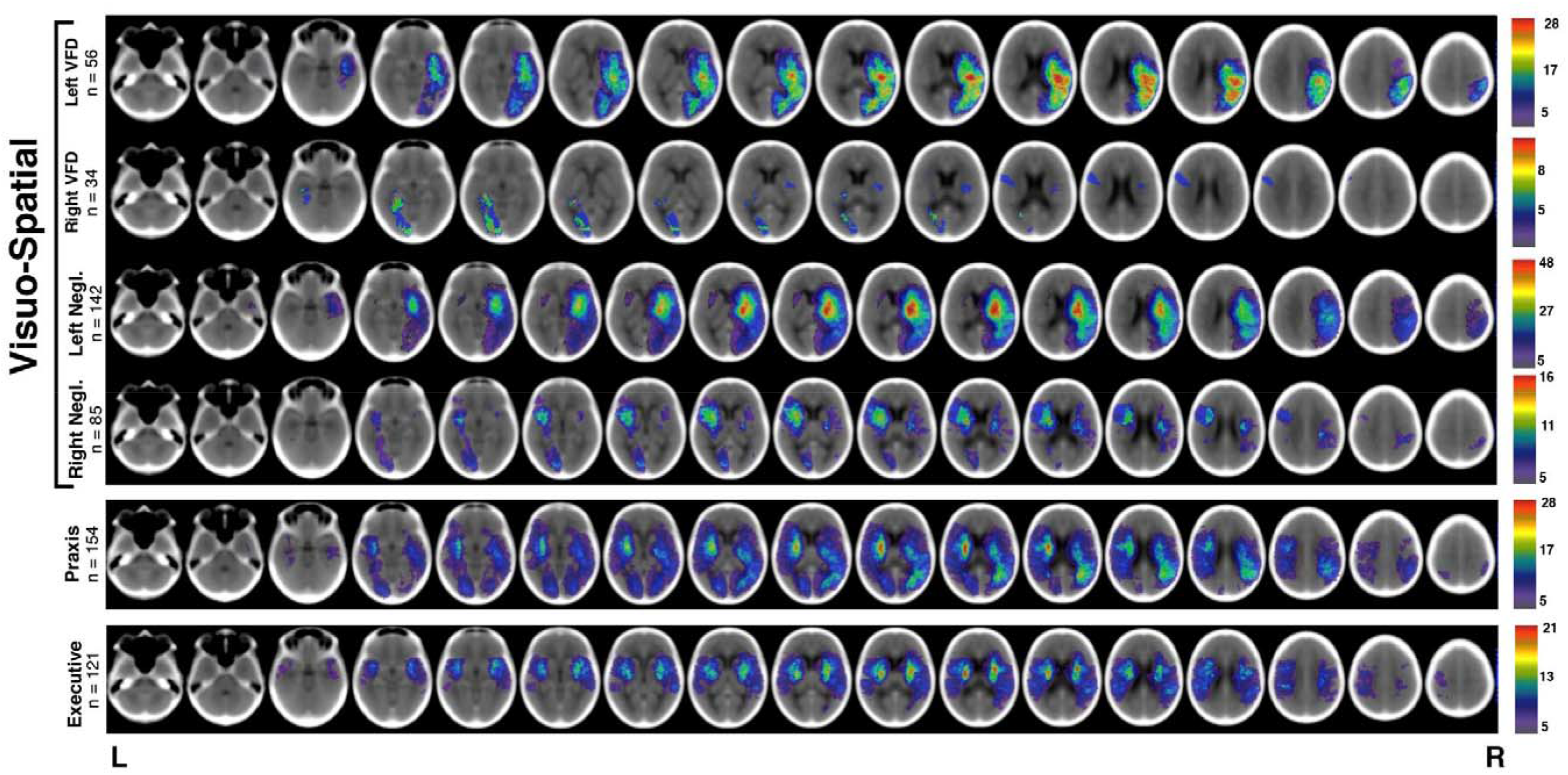
Lesion overlays for each OCS subtest. Only lesions from impaired patients are presented, with n representing the number of impaired patients. Colour indicates number of lesion overlaps within each region. Slices within the MNI z coordinates −44 to +66 are visualised

**Figure 3:**
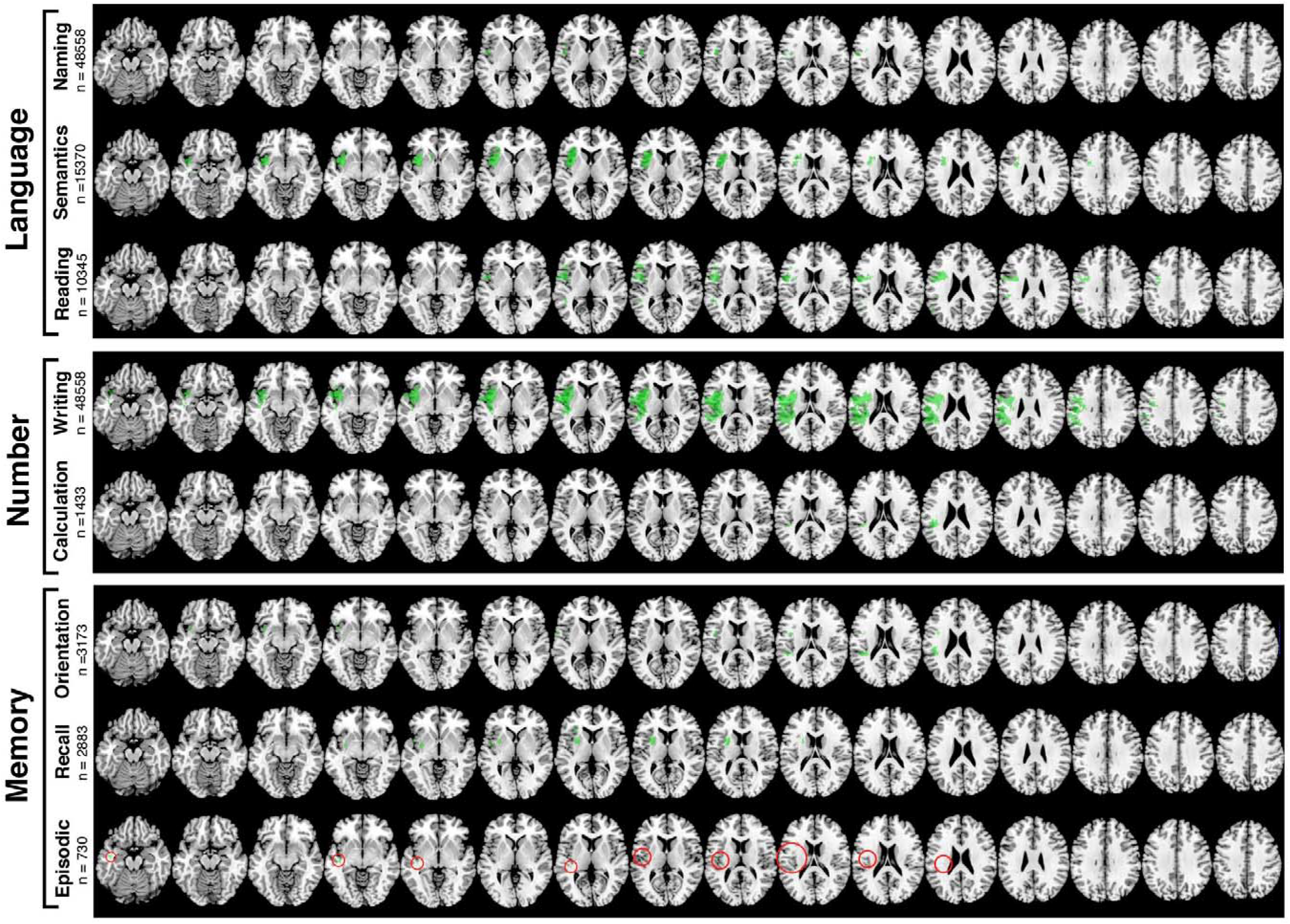
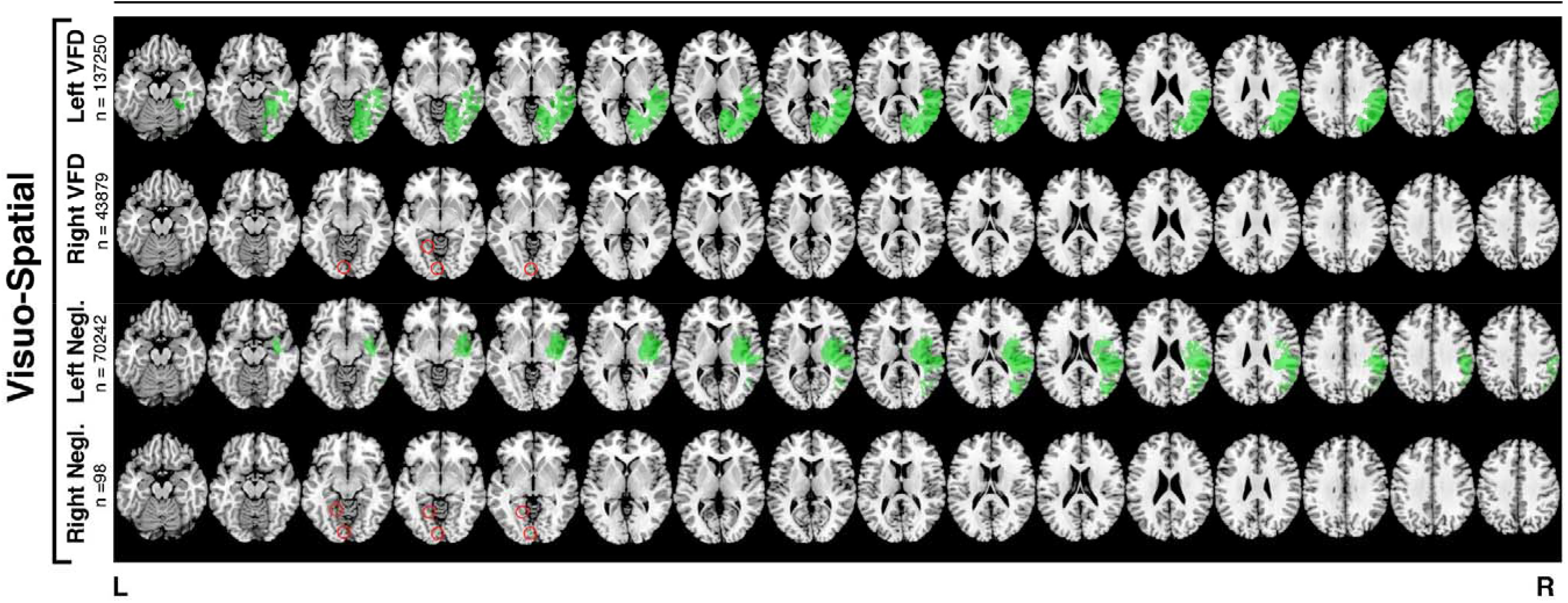
A visualisation of the voxels surviving correction for each subtest. N denotes number of significant voxels for each comparison. Small voxel clusters are highlighted in red.

Each VLSM comparison’s z-statistic ranges, significance thresholds, and peak coordinates are reported in Table 1. Table 2 provides detailed anatomical descriptions and statistics for each significant voxel cluster >50 voxels.

**Table 1:** VLSM results for each subtest analysis conducted. MNI coordinates (X,Y,Z) are provided for each analysis’ peak z-value. Z-cut denotes the z-score cut-off representing significance. This value represents a highly conservative Bonferroni correction in which the alpha level is determined by the number of voxels (N Voxels) considered in each individual analysis. Z-range represents the full range of z-values associated with all voxels analysed. N represents number of patients included and Impaired reports the number impaired in each group.

|  | Test | N | Impaired | N Voxels | z-range | z-cut | Voxels Significant | X | Y | Z |
| --- | --- | --- | --- | --- | --- | --- | --- | --- | --- | --- |
| **Language:** |  |  |  |  |  |  |  |  |  |  |
|  | Picture Naming | 543 | 178 (32.7%) | 1,167,339 | -6.39 - 2.89 | -5.355 | 1748 | -42 | 3 | 9 |
|  | Semantics | 565 | 54 (9.7%) | 1,228,158 | -7.73 - 2.14 | -5.364 | 15370 | -30 | 14 | 4 |
|  | Sentence Reading | 532 | 123 (23.1%) | 1,157,171 | -7.04 - 3.01 | -5.363 | 10,345 | -33 | 3 | 24 |
| **Number:** |  |  |  |  |  |  |  |  |  |  |
|  | Number Writing | 568 | 155 (27.3%) | 1,231,364 | -7.55 - 3.54 | -5.364 | 48,588 | -35 | 1 | 17 |
|  | Calculation | 564 | 105 (18.6%) | 1,228,868 | -6.76 - 2.99 | -5.364 | 1433 | -43 | -34 | 26 |
| **Memory:** |  |  |  |  |  |  |  |  |  |  |
|  | Orientation | 566 | 121 (21.4%) | 1,226,697 | -6.65 - 2.31 | -5.364 | 3173 | -39 | -35 | 24 |
|  | Verbal Recall | 552 | 168 (30.4%) | 1,217,657 | -7.06 - 3.43 | -5.362 | 2883 | -27 | 6 | 7 |
|  | Episodic Recognition | 559 | 141 (25.2%) | 1,224,539 | -6.32 - 3.62 | -5.363 | 730 | -43 | -12 | -6 |
| **Visuo-Spatial:** |  |  |  |  |  |  |  |  |  |  |
|  | Left Visual Field Deficit | 557 | 56 (10.1%) | 1,220,018 | -7.93 - 2.74 | -5.363 | 137,250 | 39 | -68 | 27 |
|  | Right Visual Field Deficit | 557 | 34 (6.1%) | 1,116,984 | -8.28 - 1.76 | -5.363 | 43,879 | -38 | -40 | -19 |
|  | Left Neglect | 499 | 142 (28.5%) | 1,116,984 | -7.49 - 4.00 | -5.347 | 70,242 | 43 | -33 | 25 |
|  | Right Neglect | 499 | 85 (17.0%) | 1,116,984 | -5.87 - 2.82 | -5.347 | 98 | -3 | -87 | -8 |
| **Praxis:** |  |  |  |  |  |  |  |  |  |  |
|  | Praxis | 560 | 154 (27.5%) | 1,225,043 | -5.61-3.156 | -5.363 | 4 | -17 | -84 | 1 |
| **Executive:** |  |  |  |  |  |  |  |  |  |  |
|  | Executive | 510 | 121 (23.7%) | 1,140,605 | -4.62 - 3.08 | -5.351 | 0 | -47 | -32 | 35 |

**Table 2:**
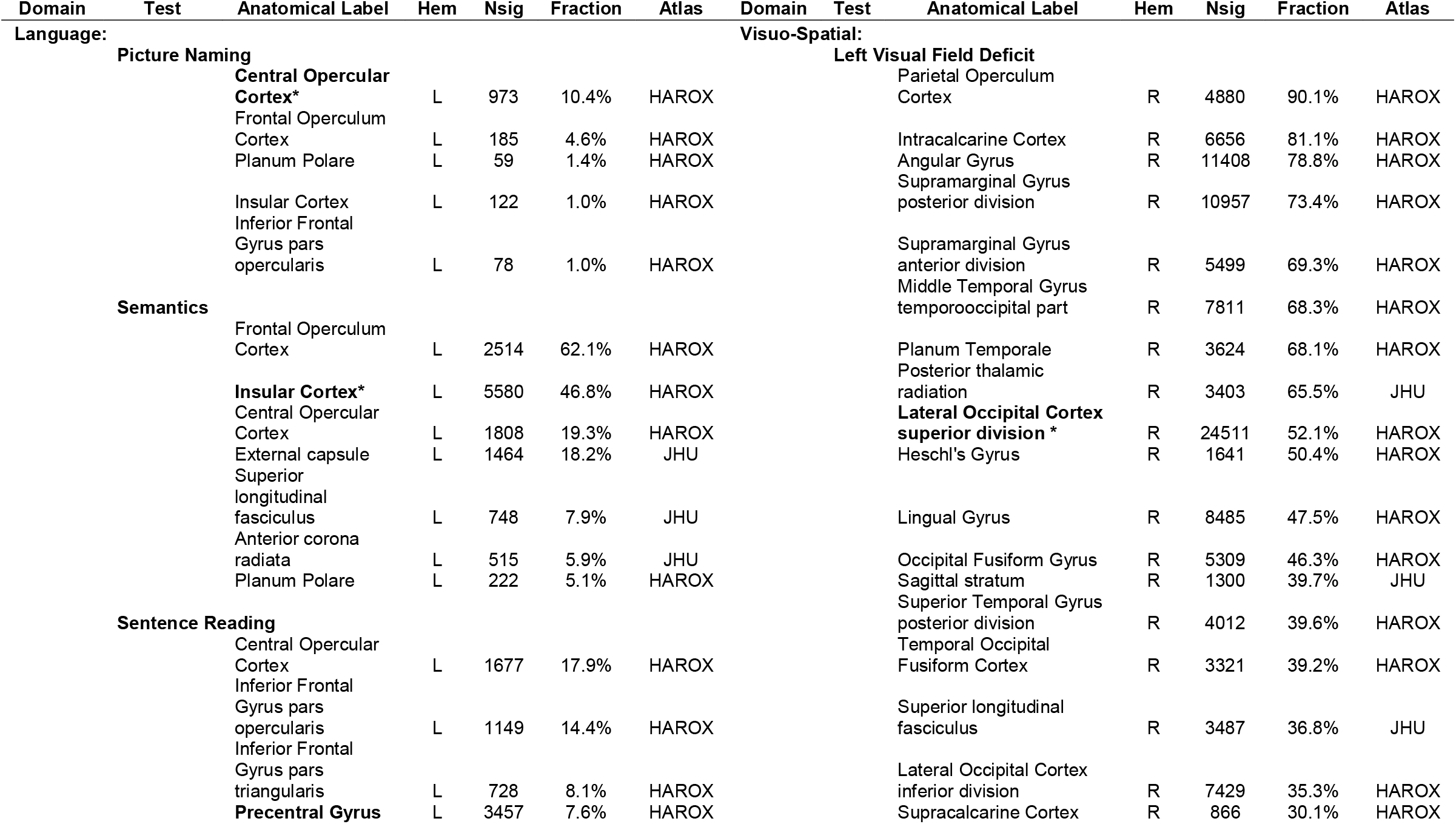
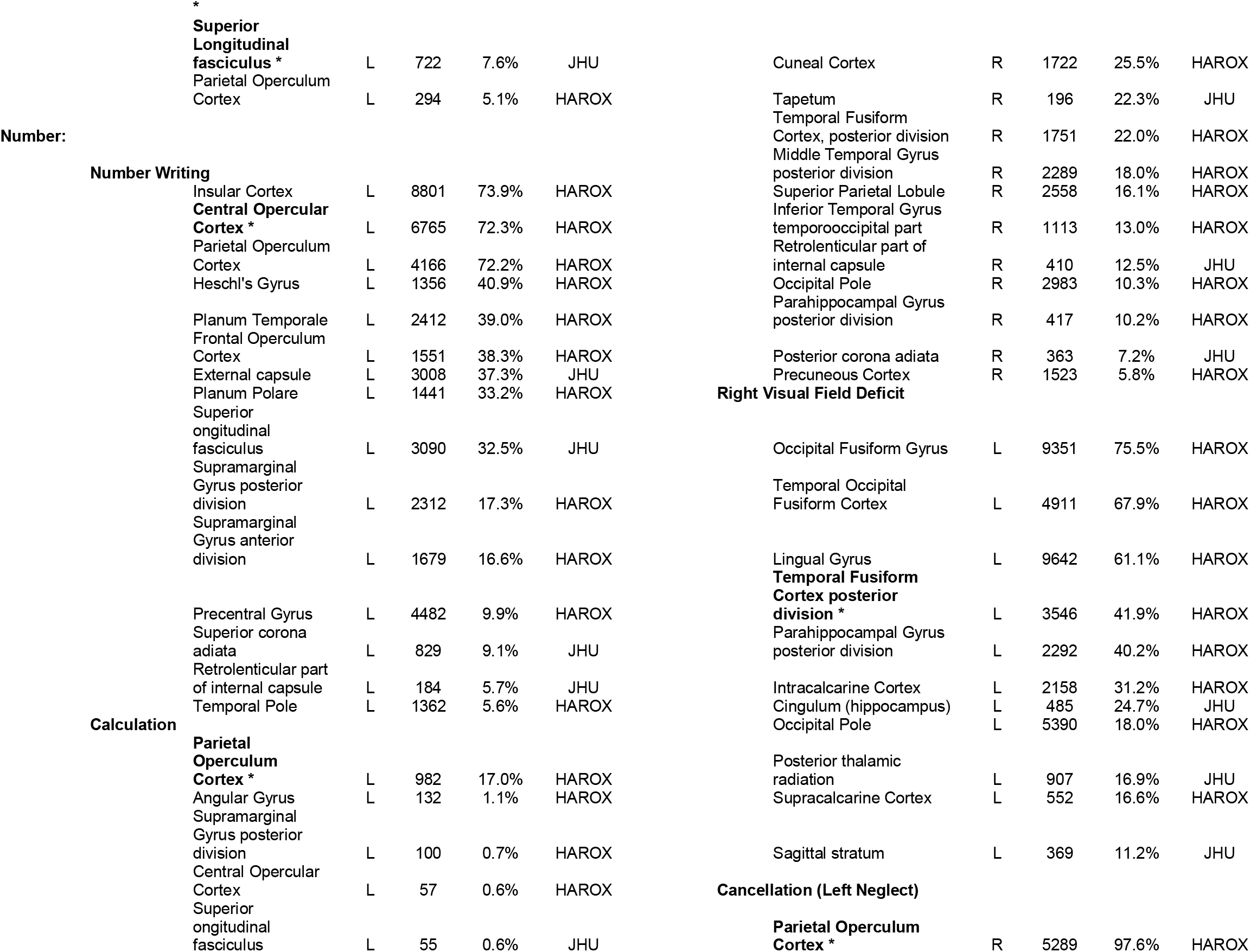
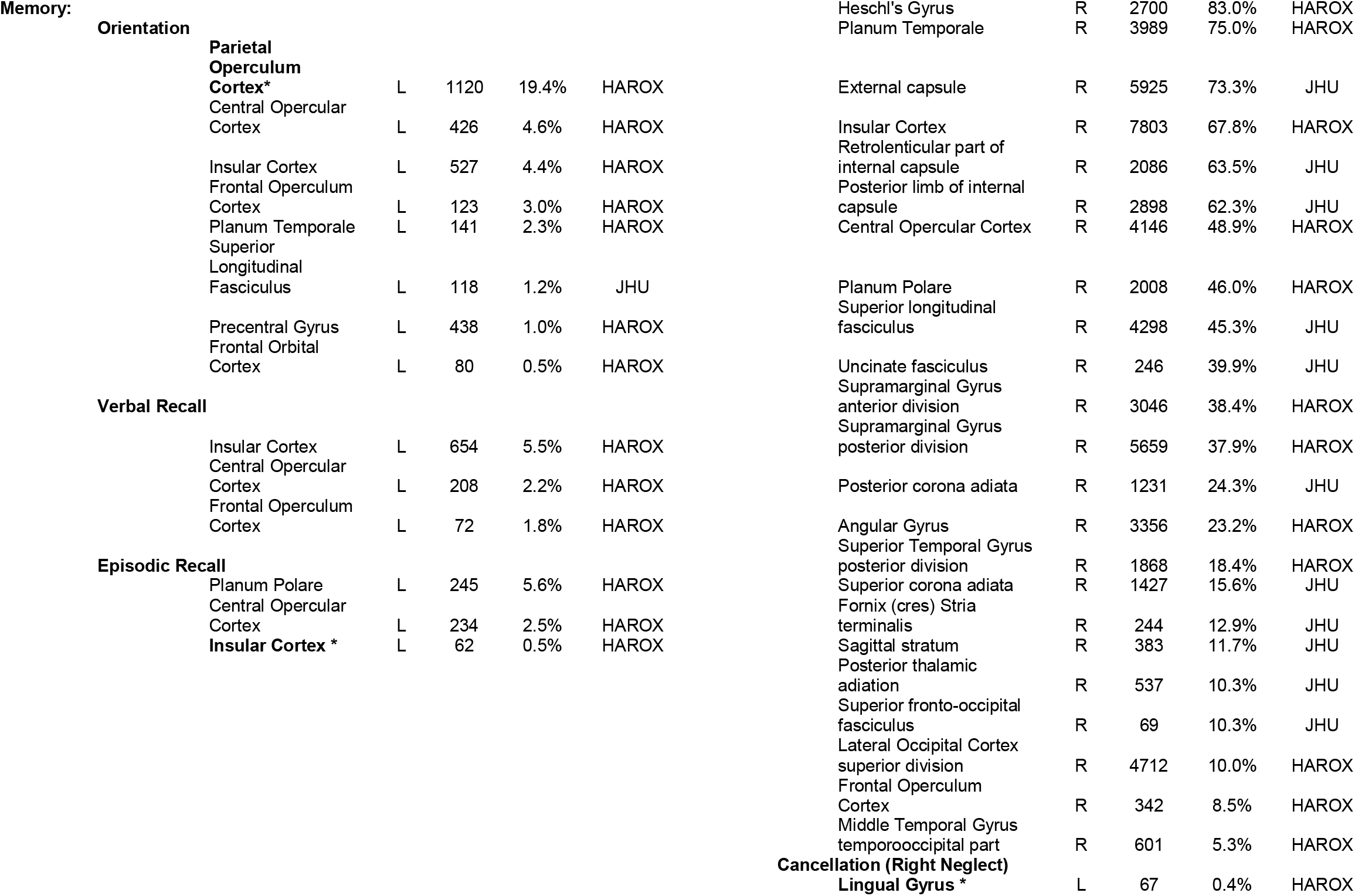
Detailed anatomical descriptions of significant voxel clusters identified in each individual analysis. Starred ROIs contain the peak z-values for each test. Only voxel clusters containing >50 voxels are reported. If a test was found to be associated with more than 5 ROIs, only ROIs containing at least 5% significant voxels are reported. Smaller clusters are included if they contained peak z-values for each test. Full anatomical descriptions for each voxel cluster are available on the Open Science Framework. Fraction represents the proportion of each ROI covered by each significant z-statistic map. All anatomical areas are defined based on the Harvard-Oxford Cortical Atlas (HAROX) and the Johns Hopkins University White Matter Atlas (JHU). Hem = hemisphere (Left/Right).

Within the language domain, VLSM analysis of the *Picture Naming Task* data indicated that language production impairment was most strongly associated with damage within the left central operculum cortex. Additional significant voxel clusters were identified within the left frontal operculum cortex and the planum polare. Receptive language impairment within the *Semantics Task* was most strongly associated with voxels within the left frontal operculum cortex, with significant areas extending into the neighbouring insular and central opercular cortices. Finally, *Sentence Reading Task* impairment was most strongly associated with damage to voxels within the left precentral gyrus and superior longitudinal fasciculus. This significant voxel cluster extended into the left central operculum cortex and inferior frontal gyrus (pars opercularis and pars triangularis).

For the domain of number ability, impairment within the *Number Writing Task* was significantly associated with voxel clusters within the left frontal-temporal area extending throughout the frontal/central operculum cortex, insular cortex, and Heschl’s gyrus and into underlying internal capsule white matter. Peak z-values for this comparison were located within the left central operculum cortex. Alternatively, *Calculation Task* impairment was most strongly associated with damage left parietal operculum cortex voxels with additional significant clusters within Heschl’s gyrus, the planum temporale, and the external capsule.

With regards to memory, *Orientation* impairment was associated with peak z-value voxels within the left parietal operculum. Additional significant voxel clusters were present within the frontal/central opercular and insular cortices. Impairment within the *Verbal Recall* task was associated with voxel clusters within the insular and central/frontal opercular cortices. The peak z-coordinates were found to correspond to the left anterior putamen.

Alternatively, *Episodic Recognition Task* impairment was most strongly associated with damage to voxels within the left insular cortex with additional clusters present within the central operculum and planum polare cortices.

Within the Visuo-Spatial domain, VLSM analysis of *Left Visual Field Task* impairment yielded a large cluster of significant voxels extending throughout the right occipital, posterior parietal, and superior parietal cortical areas and underlying white matter tracts. This large cluster primarily impacted the right parietal operculum cortex, intracalcarine cortex, and the angular gyrus. Damage to the right lateral occipital cortex (superior division) was found to be most strongly associated with impairment. Conversely, *Right Visual Field Task* impairment was found to be associated with peak z-values within the posterior division of the left temporal fusiform gyrus. Additional significant voxel clusters were located within the posterior division of the left occipital fusiform gyrus, temporal-occipital fusiform cortex, and lingual gyrus.

In comparison, analysis of left-lateralised neglect *Cancellation Task* impairment resulted in a large cluster of significant voxels extending throughout the right temporal/parietal cortices and underlying white matter. Specifically, these clusters primarily impacted the right parietal operculum, Heschl’s gyrus, planum temporale, and insular cortex. White matter tracts including the external capsule, internal capsule (retolenticular and posterior limb), and superior longitudinal fasciculus were also significantly impacted. Damage to voxels within the right parietal operculum cortex was found to be most strongly associated with left-lateralised neglect impairment. Conversely, right neglect *Cancellation Task* impairment was found to be most strongly associated with damage within the left lingual cortex.

Within the executive domain, the VLSM analysis of the Trail Making Task data yielded no significant voxels surviving correction. Finally, for the praxis domain, the *Praxis Task* impairment was associated with a very small (4 voxel) cluster centred in the left.

## Discussion

The results of this investigation provide a comprehensive validation of VLSM methods with routinely acquired clinical scans, through the demonstration of functional specialisation within the human brain. The study further extends this evidence base with an acute, clinically representative sample in the single largest cohort of domain-specific cognitive lesion mapping to date. The VLSM analyses of focal post-stroke cognitive impairments strongly suggests that distinct cognitive impairments are related to separable neural correlates and that the contributions of these correlates are necessary rather than simply correlational. The correlates identified in this study generally agree well with those found in previous investigations (discussed below), validating the use of acute CT scans for voxel lesion-mapping research. This finding is critically important, as employing clinical data can dramatically increase the scope and relevance of future VLSM analyses. This study also highlights several important strengths and limitations of employing VLSM analyses to investigate functional specialisation.

As a whole, the anatomical correlates identified in this investigation agree well with those documented in past literature. Language functions were most strongly associated with damage to left hemisphere frontal/temporal areas, agreeing well with previous research linking various language functions to similar regions. Walker et al.^49^ found that damage to the left anterior temporal lobe and middle temporal gyrus region was the best predictor of semantic error production in stroke patients with aphasia. This investigation similarly found that damage to anterior temporal areas such as the frontal operculum and insular cortex significantly predicted impairment within the OCS semantics test. Baldo et al.^50^ concluded that picture naming errors were subserved by a large network in the left peri-sylvian region including the middle temporal gyrus. Our findings are again in line with this, although our results did not specifically link middle temporal gyrus to picture naming errors, damage to a range of nearby peri-sylvian cortical and white matter areas was found to significantly predict naming impairment. Finally, Baldo et al.^51^ found that sentence-level reading impairment was predicted by damage to left inferior temporo-occipital areas including the middle/superior temporal gyri and the supramarginal gyrus. Here, we replicated these findings of left inferior temporo-occipital regions including the opercular cortex and superior longitudinal fasciculus as being significantly associated with reading impairment. In sum, the present study employing clinically applicable brief screening measures and routine clinical scans closely matches previous findings with equivalent tasks within the language domain.

In this investigation, visual field deficits were associated with damage to posterior occipital areas including the intracalcarine sulcus. This finding is in line with core visual field perception located in primary visual areas within the posterior occipital cortex^52^. The involvement of posterior occipital regions outside the primary visual cortex (e.g. lingual gyrus) can be explained by the general strong association between PCA territory strokes and visual field deficits^53^. Similarly, the significant findings outside the posterior occipital cortex (e.g. parietal operculum) can most likely be accounted for by severe visual neglect resulting in apparent visual field deficits as measured in a confrontation task. In severe cases of spatial neglect the condition is often incorrectly classified as a visual field impairment^54^.

Left visuospatial neglect was most associated with damage to a large cluster of significant voxels centred within the right temporo-parietal region^55–58^. Previous research has associated left visuospatial neglect deficits with damage to the right temporoparietal junction and superior temporal gyrus ^18,27,56,59–62^, the angular gyrus and parahippocampal region^56^, as well as the white matter tracts underlying these cortical areas (superior/inferior longitudinal fasiculi and inferior fronto-occipital fasciculi)^27,63–66^. A similar degree of heterogeneity was present within the regions found to be associated with left visuospatial neglect in this study. This variation has been traditionally accounted for by characterising visuospatial neglect as a disconnection syndrome, rather than a deficit subserved by any single neural correlate^64,67^.

Conversely, right visuospatial neglect was found to be associated with the left lingual gyrus. This finding contrasts with previous VLSM studies which have identified neural correlates for right visuospatial neglect within more anterior left hemisphere areas. For example, Suchan and Karnath^68^ found that right visuospatial neglect was subserved by the left superior/middle temporal gyri, inferior parietal lobule, and insula. Similarly, Beume et al.^69^ found that damage to the left superior and middle temporal gyrus, temporal pole, frontal operculum, and insula were the strongest predictors of right neglect. However, previous research has identified a wide range of cortical and subcortical right hemisphere regions which are associated with left neglect^61,69^. It is highly plausible that similar variance exists within the left hemisphere areas responsible for distributing attention across space. Additional research is needed to further characterise any potential interactions between different left hemisphere cortical areas involved in spatial neglect.

Notably, a VLSM study by Moore et al.^70^ considering a subset of the patient data reported here identified a correlate of right allocentric neglect within the left parahippocampal gyrus, but found that right egocentric neglect was significantly associated with exclusively ipsilateral damage to the right parietal operculum. This result likely differed from the present study due to the differentiation between egocentric and allocentric neglect impairment. The present investigation did not aim to investigate differences between egocentric and allocentric neglect, but instead compared patients with and without any type of right-lateralised neglect deficit.

Not all cognitive deficits included in this investigation were found to be associated with significant clusters of localised voxels. While VLSM analysis of praxis impairment did yield a cluster of four significant voxels within the left intracalcarine cortex, this voxel cluster was not large enough to be included within this investigation’s analysis. Previous research has strongly suggested that praxis functions are not subserved by a single, localised neural correlate but instead represent diffuse, network-based cognitive functions^71,72^. For example, Hoeren et al.^71^ found that gesture imitations were associated with damage to the left lateral occipito-temporal, posterior inferior parietal lobule, posterior intraparietal sulcus, intraparietal sulcus, and superior parietal lobule. However, in order to successfully imitate gestures participants must first accurately perceive the presented stimulus. This reliance on visual information suggests that visual perception areas also play a critical role in supporting praxis abilities. Visual examination of the lesion overlays of participants with praxis impairment demonstrates a clear, bilateral damage pattern associated with this disorder. Given that VLSM is less well suited to identifying the neural correlates of bilateral functions^30^, additional analyses are needed to more thoroughly understand the neural correlates of praxis impairment.

Similarly, impairment on the OCS executive function task was not found to be significantly associated with damage to any voxels in VLSM analysis. While executive functions such as response inhibition, goal-directed behaviour, and self-monitoring are classically linked to damage within the left or right pre-frontal cortex^20,73,74^, various components of executive function have been associated with damage to a wide range of neural correlates. For example, Varjačić et al.^22^ found that the left insula plays a critical role in subserving executive set switching. Similarly, specific executive function components, such as inhibition, have been found to be associated with distinct neural regions even within the prefrontal cortex^74^. This diversity is clearly illustrated within this study’s lesion overlay of patients exhibiting executive function impairment, as bilateral lesions involving a range of primarily the white matter underlying temporo-parietal regions was found to be impacted. In sum, the existing literature strongly suggests that executive function, like praxis, is best understood as a diffuse function subserved by multiple, interconnected neural correlates.

VLSM is designed to identify localised neural correlates and is a less appropriate method for identifying the underlying mechanisms of diffuse functions or disconnection syndromes^75^. When any single voxel is analysed in VLSM, the behavioural scores of all patients with lesions to the relevant voxel are compared to all patients without damage at that voxel, regardless of behavioural phenotype. If a specific deficit is supported by multiple or bilateral correlates, this approach can lead to the impact of lesions at different critical sites cancelling each other out, leaving only a single significant correlate^30,31^. This implies that this study’s failure to identify the neural correlates of praxis and executive function is not indicative of insufficient data, but rather illustrates one of the established weaknesses within VLSM methodology. Alternative, network-based lesion mapping techniques are more appropriate for investigating the neural correlates of disconnection syndrome cognitive deficits such as executive function.

The inclusion of routine imaging rather than research-specific scans offers an exceptionally valuable resource for improving the scope and quality of research aiming to identify the neural correlates of behaviour. Although individual clinical CTs may be likely to report false negatives, these scans are extremely unlikely to yield false positives in VLSM analyses, making them a valuable resource for future research. These results also demonstrate the utility of standardised post-stroke cognitive assessment for detecting and differentiating between distinct patterns of cognitive impairments. The OCS bedside assessment alone was able to reliably identify and differentiate post-stroke cognitive deficits, teasing apart distinct patterns of underlying damage^25,26^. These findings further validate the use of this screening tool as an effective method for identifying domain-specific rather than domain-general deficits^26^. The consistent involvement of classical language areas across the number and memory domains does imply that these tasks, though inclusive for patients with language impairments, tend to involve a degree of language dependency. Nevertheless, the analyses demonstrated that cognitive functions such as verbal recall and episodic recognition, were related to distinct neural correlates within the memory domain.

Similarly, the present analysis was able to distinguish neural correlates of distinct functions which often present similarly in behaviour. For example, left visual neglect and left visual field deficits were found to be associated with distinct impairment patterns. Even so, when visuo-spatial neglect is severe, the attentional orientation to the neglected field can be so severely affected that patients fail a confrontation test designed to detect a perceptual deficit. This was apparent also in the lesion correlates for the confrontation visual field task which showed involvement of more anterior (non-visual) temporo-parietal areas. This is in line with previous findings that severe visuospatial neglect is commonly misdiagnosed as a visual impairment^54^.

The results of this investigation also highlight some inherent limitations associated with using VLSM techniques. First, regardless of the scan type used, there will inevitably be some degree of measurement error associated with creating quantitative lesion masks from uncertain clinical data. Investigations including low numbers may produce results which are skewed by this noise. Future VLSM analyses must therefore aim to include large, representative samples with sufficient lesion coverage to detect statistically significant, generalisable effects. These large group analyses do require strict corrections for multiple comparisons inherent in VLSM techniques. Second, while VLSM can effectively identify the correlates associated with localised functions, this technique is not designed to identify networks of multiple areas contributing to more distributed cognitive abilities. Researchers should use extreme caution when using VLSM to investigate potential disconnection syndromes.

### Study Limitations

Ideally, domain-specific cognitive impairments should be diagnosed based on agreement between multiple, independent assessments^26,76,77^. Given this investigation’s reliance on existing data, it was not possible to include data from additional tests. Additionally, different underlying problems can result in impairment on any single task. For example, visually impaired patients would be expected to fail a reading task for very different reasons than a patient with language impairments. Such variation likely induced some degree of noise into this investigation’s analyses, though the large sample size likely prevented this inherent noise from significantly impacting anatomical results.

While this study’s results indicate that separable lesion patterns are associated with different cognitive impairment profiles, we emphasise that this is mainly a research advance and that any potential clinical utility of using this to provide a neuroimaging-based diagnosis of cognitive difficulties is extremely limited. First, clinical CT imaging has a high-false negative rate^39^. Second, a diverse range of lesions both within and outside identified critical lesion sites can cause a specific cognitive impairment. This problem is particularly apparent in disconnection syndromes, such as visuospatial neglect and executive function disorders ^20,64,67,73^. Finally, standardised neuropsychological testing represents a simpler and more reliable method for identifying cognitive impairments^24^. The findings presented in this investigation are meant to illustrate large-scale patterns of functional specialisation based on routinely collected data, not to guide diagnosis of cognitive impairments in clinical settings.

## Conclusions

We demonstrated the validity of using routinely collected clinical neuroimaging data for VLSM analysis with standardised cognitive screening data. A domain-specific bedside assessment alone was able to reliably identify and differentiate post-stroke cognitive deficits, teasing apart distinct patterns of underlying damage, confirming findings of focal stroke lesions impacting distinct cognitive domain functioning post stroke. The present demonstration opens up VLSM techniques to a wealth of clinically relevant lesion mapping studies to capitalise on using existing clinical brain imaging.

## Data Availability

All behavioural summary data and analysis output files are openly available on the Open Science Framework47 (hhttps://osf.io/dc7ay/). All additional data is available upon request.

## Acknowledgements

We would like to express our sincere gratitude and admiration to the late Prof Glyn W Humphreys, who initiated the OCS work and led the OCS-Care study. The OCS study was supported by the National Institute for Health Research Clinical Research Network. We nwould like to thank Jacob Levenstein and Anja Varjacic for their contributions to pre-processing the lesion data employed in this investigation. We also acknowledge the contributions to data collection and curation for the OCS data made by Ms Ellie Slavkova, Ms Grace Chiu, and Ms helpful feedback on the final manuscript.

## Author Contributions

MJM designed and conducted analysis and drafted the manuscript. ND conceptualised the study, supervised analysis, and edited manuscript drafts.

